# Implementing a Telemedicine Curriculum for Internal Medicine Residents during a Pandemic: The Cleveland Clinic Experience

**DOI:** 10.1101/2020.10.12.20211136

**Authors:** David J. Savage, Omar Gutierrez, Bryce Montane, Achintya Dinesh Singh, Eric Yudelevich, Jamal Mahar, Andrei Brateanu, Lakshmi Khatri, Catherine Fleisher, Stacey Jolly

**Affiliations:** Resident, Internal Medicine Residency Program, Cleveland Clinic Foundation; Associate Program Director, Internal Medicine Residency Program, Cleveland Clinic Foundation; Director of Ambulatory Curriculum, Internal Medicine Residency Program, Cleveland Clinic Foundation

## Abstract

**Introduction:** Telemedicine is an important element of healthcare. However, until the COVID-19 pandemic, training in telemedicine was not a substantial element of most residency programs. Social distancing measures changed this. The Cleveland Clinic Internal Medicine Residency Program (IMRP) is one of the largest programs in the United States, which made the task of developing and adopting an effective, expedited telemedicine curriculum challenging. Our goal was to implement a system for teaching telemedicine care skills and supervising the care provided by residents during virtual visits.

**Methods:** This study was started in April 2020. We developed and implemented a resident-led curriculum and training program for providing telemedicine care in less than five weeks. This entailed creating a formal training program for residents, creating a resource guide for the different video communication tools, and training preceptors to safely supervise care in this new paradigm. We also created an assessment instrument in our education evaluation system that allows residents to receive feedback on their performance during virtual appointments.

**Results:** Over 2000 virtual visits were performed by residents in a span of 10 weeks. Of 148 residents, 38% responded to the pre-participation survey. A majority had no prior telemedicine experience and expressed only slight comfort with the modality.

**Discussion:** Through collaboration with experienced residents and faculty, we expeditiously developed an enhancement to our ambulatory care curriculum to teach residents how to provide virtual care and help faculty with supervision. We share our insights on this experience for other residency programs to utilize.

## Introduction

The first documented cases of COVID-19 in Ohio occurred on March 9, 2020^1^. One day later, the Cleveland Clinic’s leadership announced that all in-person meetings were cancelled and ambulatory patient visits would be virtual. This was a major paradigm shift for our Internal Medicine Residency Program (IMRP), where one-third of training time is spent in ambulatory clinics. Teaching residents at one of the largest programs in the country to provide virtual visits on short notice was an enormous endeavor. In addition, we needed to make sure that all faculty had the tools they needed to precept residents in a completely virtual format.

A cohort of Cleveland Clinic faculty physicians have been performing virtual patient visits since 2014 using the American Well platform^2^. They conducted 41,000 virtual visits in 2019 and over 100,000 such visits by February 2020^3^. Residents, however, had not been trained in delivering virtual care prior to the COVID-19 pandemic. In any given week, our IMRP residents complete around 840 ambulatory patient encounters. The avoidance of in-person visits for routine care as a result of social distancing measures would have deprived our community of considerable primary care, as well as limited learning opportunities for residents.

Our IMRP’s clinician-educator track (CET) residents began building a telemedicine curriculum before the start of the pandemic. The original plan was for a slow, stepwise roll-out of the curriculum with the new interns in Fall 2020. However, the urgent need created by the pandemic called for an accelerated implementation of this nascent telemedicine curriculum.

## Methods

The Accreditation Council for Graduate Medical Education (ACGME) had planned to integrate telemedicine into the Common Program Requirements in July 2020. Considering the pandemic, precepted telemedicine encounters were permissible immediately^4^. Our telemedicine curriculum allowed us to quickly train residents and preceptors to serve our community with virtual visits. Study of the telemedicine curriculum was started in April 2020 and was determined to be exempt by the Cleveland Clinic IRB.

Our ambulatory sites accommodate 148 residents and vary in size and target demographics. Residents function as primary care physicians during their clinic week (4+1 model) in nine different clinic sites. Implementing this program involved preparing and standardizing clinic work-flow documents for residents and a precepting guide for faculty across the different ambulatory sites across the different ambulatory sites (see Appendices). We created an orientation presentation to standardize virtual visits inspired by external resources from the American College of Physicians (ACP), the Ohio State Medical Association (OSMA), and the ACGME. Telemedicine training for residents was conducted virtually by Zoom, a practice that has been widely used at other institutions to implement virtual curricula^5^.

### 1. Trainee Pre- and Post-Participation Surveys

Residents were surveyed about their prior telemedicine experience and perceptions of telemedicine before participating in this telemedicine curriculum. We then surveyed residents at the end of the academic year to assess their perception of the curriculum in improving their capability to provide virtual primary care.

### 2. Communication Competencies

We considered it paramount to teach our residents how to communicate and perform remote exams with virtual tools, which require unique communication skills^6^. Residents watched a recording of a simulated encounter prior to their first real encounter (see Appendices). We demonstrated potential pitfalls of a virtual encounter (from technology-specific issues such as audio or connectivity problems, to challenges in virtual verbal communication) and paused periodically for discussion. This gave residents an opportunity to share their impressions with the group and brainstorm possible solutions. The *Cleveland Clinic Communication Tips for Virtual Visits* (see Appendices) was used to teach resident essential skills (e.g. conveying respect, setting an agenda, and providing reflective listening using non-verbal communication). The presentation also reviewed practical tips such as looking at the camera instead of the screen, which provides the appearance of direct eye contact. Initially, virtual visits were conducted in dedicated patient rooms on campus, but eventually many visits were conducted remotely in order to provide appropriate social distancing for trainees and staff. Medical decisions were always discussed with and approved by attending physicians, either over the video platform, by phone, or in person.

### 3. Virtual Physical Examination Competency

Physical examinations in telemedicine must be augmented to fit the virtual nature of the encounter. Two inherent challenges with virtual visits are the physician’s ability to describe the exam he/she needs, and the patient’s ability to perform the exam on video. In our program, we taught the virtual physical exam with Zoom. We used problem-based questions and interactive cases, with symptoms ranging from ankle pain to vision changes. During these scenarios, we emphasized creativity. We showed residents how to teach a patient the Ottawa ankle rules to address ankle pain, for example, thus saving them a visit to the emergency department.

### 4. Telemedicine Technology Platform Competency

Virtual encounters were initially offered using a variety of platforms including American Well, Doximity, Skype, and Apple FaceTime. We created a resource slide deck with practical tips on how to install and use each of those platforms (see Appendices). One month later, our institution started using a secure version of Zoom integrated into our EMR (Epic Systems) to streamline virtual visits. Our slide deck was updated to incorporate this new platform. Residents also viewed an institutional video that walked them through using Zoom within Epic for virtual visits.

### 5. Teaching Documentation of Virtual Encounters

Residents and preceptors were taught how to appropriately document virtual encounters. New documentation elements to the standard ambulatory note included a statement that the encounter was conducted by phone or video, an estimate of the time it took to provide care, and a description of the findings of the virtual physical exam, if performed. Preceptors also needed to attest that the visit was provided either by phone or video, and indicate whether they were present for a portion of the encounter. These documentation changes were facilitated with new templated phrases in Epic that all residents and staff physicians could use.

### 6. Virtual Encounter Formative Trainee Evaluation

We created a formal evaluation process to help residents improve their practice. Residents were still required to precept their patient care with a staff physician. Staff physician could join the encounter at any time, especially during the visit wrap-up where the resident covers the plan with the patient and answers questions. Residents were also able to get direct observation and feedback during collection of the patient’s history and their performance of the physical examination in the virtual setting. This mirrored the process for in-person visits. We created a mini-clinical evaluation exercise (Mini-CEX) for telemedicine centered around the ACGME core competencies^7^. Each resident was encouraged to send an electronic mini-CEX survey to faculty to get actionable feedback on their virtual visit, and CEX evaluations were geared to evaluate for the nuances of the virtual encounter.

### 7. Precepting the Virtual Encounter

Dynamic changes in recommendations for trainee virtual visit by the ACGME and billing and supervision rules from the Centers for Medicare & Medicaid Services (CMS), made regular updates for all provider participating in virtual care precepting environment paramount. Our precepting team clinic leaders collaborated to identify champions for maintaining updates to ACGME and CMS rules and providing weekly and as-needed updates to all faculty, residents, and support staff involved in scheduling. This incorporated changes to workflow for consent to encounter, location of preceptor (direct or indirect supervision) and identification of the types of encounters for which the CMS primary care exception, where the preceptor does not interact with the patient, could be utilized during the COVID-19 public health emergency. Although the cadence of these changes in the future is anticipated to be slower, the successful utilization of a telemedicine curriculum includes a support structure to update and incorporate rapid changes as needed.

## Results

Our team designed and implemented the telemedicine curriculum within four weeks. All 148 ambulatory residents in our IMRP have now been trained in performing virtual visits and have completed more than 2000 such visits since early April 2020. In total, 56 residents completed the orientation pre-participation survey with a total response rate of 38%. Forty-three residents completed the post-participation survey, with a total response rate of 29%. Of residents responding on the pre-participation survey, 68% had no prior telemedicine experience and indicated limited comfort with the modality (Table 1). The post-participation survey assessed the residents’ perception of the quality of this curriculum in teaching telemedicine skills. A majority of respondents were third-year residents (n = 20, 46.5%), and 67.4% of respondents (n = 29) reported learning new knowledge about telemedicine because of the curriculum that was offered (Table 2).

**Table 1.** Resident telemedicine orientation pre-participation survey responses (N = 56)

| Responses |  | Respondents with prior telehealth experience |  | Respondents comfort with conducting telehealth visits |  |
| --- | --- | --- | --- | --- | --- |
| Current PGY year | % |  |  |  |  |
| PGY-1 | 46% (n=26) | Medical school | 2% | Very comfortable | 7% |
| PGY-2 | 33% (n=19) | Demonstration by attending | 23% | Comfortable | 23% |
| PGY-3 | 19% (n=11) | None to date | 68% | Neutral | 39% |
|  |  | Other | 7% | Uncomfortable | 29% |
|  |  |  |  | Very uncomfortable | 2% |
|  |  | Respondents view of benefits of telemedicine |  | Respondents who expect to integrate telemedicine into future practice |  |
|  |  | Improved patient outcomes | 21% | Strongly agree | 36% |
|  |  | stronger physician-patient relationship | 39% | Agree | 36% |
|  |  | access to data (e.g. home measuremen | 43% | Neutral | 18% |
|  |  | improved efficiency | 63% | Disagree | 9% |
|  |  | better access for patients | 80% | Strongly disagree | 2% |
|  |  | Respondents view of barriers to using telemedicine |  |  |  |
|  |  | Providers lack of familiarity with the technology | 55% |  |  |
|  |  | Weaker physician-patient relationship | 46% |  |  |
|  |  | Too difficult for my patients to use | 63% |  |  |
|  |  | Lack of time to implement care | 11% |  |  |
|  |  | Privacy concerns | 18% |  |  |
|  |  | Reimbursement concerns | 38% |  |  |
|  |  | Documentation concerns | 29% |  |  |
|  |  | Other: lack of full physical exam | 14% |  |  |

**Table 2.** Resident post-participation survey responses 12 weeks after starting the telemedicine curriculum (N = 43)

| Responses |  | Overall, I was satisfied with this learning activity. |  | I learned new knowledge and skills from the telemedicine curriculum. |  | I found the material in this learning activity to be relevant and up-to-date. |  |
| --- | --- | --- | --- | --- | --- | --- | --- |
| Current PGY year | % | Strongly Agree | 25.6% | Strongly Agree | 18.6% | Strongly Agree | 32.6% |
| PGY-1 | 25.6% (n=11) | Agree | 44.2% | Agree | 48.8% | Agree | 51.2% |
| PGY-2 | 27.9% (n=12) | Neutral | 23.3% | Neutral | 27.9% | Neutral | 14.0% |
| PGY-3 | 46.5% (n=20) | Disagree | 4.7% | Disagree | 2.3% | Disagree | 0.0% |
|  |  | Strongly Disagree | 2.3% | Strongly Disagree | 2.3% | Strongly Disagree | 2.3% |
|  |  | I would recommend this learning activity to others. |  | I will be able to apply the knowledge and skills learned for future encounters. |  | The content was relevant to my job-related needs. |  |
|  |  | Strongly Agree | 27.9% | Strongly Agree | 30.2% | Strongly Agree | 32.6% |
|  |  | Agree | 44.2% | Agree | 53.5% | Agree | 55.8% |
|  |  | Neutral | 20.9% | Neutral | 11.6% | Neutral | 7.0% |
|  |  | Disagree | 4.7% | Disagree | 2.3% | Disagree | 4.7% |
|  |  | Strongly Disagree | 2.3% | Strongly Disagree | 2.3% | Strongly Disagree | 0.0% |
|  |  | The learning activities and/or materials were effective in helping me learn the content. |  | The scope of the material was appropriate for my needs. |  |  |  |
|  |  | Strongly Agree | 21.4% | Strongly Agree | 25.6% |  |  |
|  |  | Agree | 57.1% | Agree | 55.8% |  |  |
|  |  | Neutral | 16.7% | Neutral | 11.6% |  |  |
|  |  | Disagree | 4.8% | Disagree | 4.7% |  |  |
|  |  | Strongly Disagree | 0.0% | Strongly Disagree | 2.3% |  |  |

## Discussion

Our experience demonstrates that teaching and precepting telemedicine in resident ambulatory clinics is feasible. Implementing any type of curriculum or training program requires extensive preparation, involvement of multiple stakeholders, and competing time constraints. The urgency of the COVID-19 pandemic and the resulting changes to healthcare delivery allowed us to move this process along quickly within a major tertiary medical center and the one of the largest internal medicine residency programs in the United States. This project is also notable for having significant trainee leadership from inception.

In many regards, the process for scheduling, performing, precepting, and documenting a virtual visit parallels a traditional encounter. One key difference, however, is the physical exam. Each encounter started as a phone call, and then video could be added if needed. Many of our older patients did not have a video-enabled device, which made visual appointments challenging, even when warranted. It was also cumbersome at first for residents to find a video platform that would work for each patient. Patients were much more likely to adopt video calls early on when we offered a wide variety of choices. Our institution’s rollout of Epic with embedded Zoom video made visual appointments more uniform.

To evaluate our curriculum, we used pre- and post-participation surveys modeled after the work of others in creating a longitudinal telemedicine curriculum^8^. The pre-participation results indicated that a majority of residents had not had prior telemedicine experience, but that they felt it was an important skill to learn and would be part of their future practice (Table 1). The post-participation survey indicated that this curriculum was perceived as effective at teaching telemedicine skills that could be applied immediately to serve patients (Table 2). Residents selected Strongly Agree or Agree to each of the eight statements on the post-participation by over 60% per question, suggesting that the curriculum provided benefit in teaching telemedicine skills. Free response survey answers provided both positive and growth-oriented feedback. In regard to the content, many residents felt that it helped bridge the skill gap created by the pandemic. One respondent wrote “It was very helpful as I had no formal training in telemedicine before this time and I had to do virtual visits in my Resident PCP clinic during COVID-19.” Regarding the delivery of the content, some respondents wrote that the classroom course was “not interactive enough” or that “too much of the presentation was obvious or common sense.” Thus, while beneficial, there remains significant room for improvement in the telemedicine curriculum by building upon this feedback in coming years.

Now that our residents have had at least three ambulatory weeks of telemedicine encounters, our next step will be to assess their self-reported competency in providing telemedicine care by phone or video. Since a majority of residents from the 2019-2020 academic year did not have prior telemedicine experience, it will be helpful to understand how residents’ skillsets have changed because of this curriculum and hands on experience in clinic. In addition, residents are now soliciting formal objective feedback from their ambulatory preceptors on their performance with quarterly summative evaluations and on-demand Mini-CEX evaluations specific to virtual visits. These feedback tools will help learners improve and provide a further assessment of this curriculum. The new interns in 2020-2021 will offer in-person visits exclusively for the first six months of the year, and then will have at least one telemedicine slot per ambulatory week thereafter. We believe this will allow the newest trainees time to better understand the workflow at our institution and the tools available to serve their patients in a hands-on, supervised environment before those interns begin performing telemedicine encounters. They will also receive the Introduction to Telemedicine course that was offered to all residents in our residency program at the start of the pandemic.

### Limitations

Our limitations were a low survey response rate and variable preceptor experience with telemedicine. The response rate for the pre-participation survey was higher than the post-participation survey, but the response rate on both instruments was low. This is likely because of the accelerated pace that this curriculum was rolled out to meet the needs of our learners and patients. The pre-participation survey was offered immediately before the telemedicine curriculum was taught, while the post-participation survey was distributed electronically at the end of the academic year. We had the most success with getting pre-participation surveys completed by providing time at the start of the didactic course for residents to fill them out with their phones. We also kept the number of survey questions short to encourage participation. It has thus far been even more challenging to get learner post-participation feedback because of their busy work schedules and the ongoing pandemic. The post-participation survey response rate was also impacted by timing. It was distributed at the end of the academic year when many residents were preparing to graduate or advance to the next training year. Anecdotally, though, residents have told their preceptors and program leaders that this curriculum made them feel more comfortable with providing telemedicine care.

Lack of preceptor experience with ambulatory telemedicine was also a limitation early on, but this improved with time, practice, and the creation of a faculty preceptor guide. Since preceptors supervise resident care, the biggest change in workflow has been at the end of each encounter. If a resident wants the preceptor to examine the patient or be available on the line to listen to the wrap-up with the patient, the preceptors need to either join the resident in person or connect to the encounter by three-way video or phone. This was something that our faculty mentors and learners adapted to quickly, and this would be a very manageable barrier within any other residency program, especially those of smaller size. Despite challenges in assessment and ambulatory precepting, this curriculum has served the needs of our residents well, and it will be a sustainable element of our training program.

The ideal telemedicine curriculum for residents should provide training in how to conduct a virtual interview and exam, as well as mechanisms for receiving feedback from precepted virtual visits. Patient access to video-enabled technology is a limitation, but phone visits can be effective when a physical exam is not warranted. We hope our experience can guide other programs in developing a lasting telemedicine curriculum that prepares physicians-in-training for a practice landscape that includes virtual visits.

#### What is already known on the subject

- Prior to March 2020, telemedicine training for residents and fellows was limited, even though telemedicine was quickly emerging as widespread tool for providing care
- Telemedicine curricula for residents and fellows are quite new, but their development was accelerated by the COVID-19 pandemic

#### Study’s Main Messages

- A standardized curriculum for trainees and faculty members can build competency with providing care using telemedicine
- Trainee leadership in building a telemedicine curriculum can accelerate development and increase buy-in from both faculty and peers.

## Supporting information

Communication Tips for Virtual Visits at CCF

Precepting Guide for Telemedicine at CCF

Telemedicine Training Video

Telemedicine Survey - Pre-Participation

Telemedicine Survey - Post-Participation

## Data Availability

All original data is available upon request.

## Appendices

A. Communication Tips Virtual Visits CCF.pdf
B. Telemedicine Training Video.mp4
C. Telemedicine Survey – Pre-Participation.pdf
D. Telemedicine Survey – Post-Participation.pdf
E. Precepting Guide for Telemedicine CCF.docx

