## Supplementary material for "Implementing a Telemedicine Curriculum for Internal Medicine Residents during a Pandemic: The Cleveland Clinic Experience": Communication Tips for Virtual Visits at CCF

Cleveland Clinic
Center for Excellence in Healthcare Communication

### Top 10 Tips for Virtual Visits Clinician Communication

**The following are 10 best practices based on the R.E.D.E. Model® of healthcare communication for communicating effectively with patients in a virtual visit.**

**The Center for Excellence in Healthcare Communication (CEHC) provides evidence-based training and resources in healthcare communication and service excellence throughout the enterprise. For more information or to connect with CEHC**

#### 1. Convey Value and Respect with your welcome

**Why?** When patients feel like you see them as a person, you create a safe space that invites their trust and a more open exchange of information.

**What?** Brief behaviors (e.g., smile, look at the camera versus the screen to simulate eye contact, gather names from everyone on camera) at the start of a visit demonstrate our capacity to see patients as people. Acknowledge the virtual nature of the interaction.

**How?** *"Hello Mr/s. ____. Thank you for inviting me into your home so that we can have a conversation. It is good to see you again. It is wonderful that your wife can join us. How have you been since our last visit together?"*

#### 2. Introduce the Technology

**Why?** Orient patients to the benefits of a virtual visit as well as the difference from an in person visit. Such an orientation helps set realistic expectations thereby reducing any possible annoyance that may be associated with the use of technology.

**What?** Identify the technology

**How?** *"I'd like to talk briefly about what it's like to have a virtual visit. As you already found out, your home is more comfortable than an office waiting room. But, since this isn't in person, please know that if it seems like I'm not looking at you, that's probably because I'm looking at you on the screen. You should also know that I have a computer here with your medical records and I may be looking at that periodically. I will type a few notes during the visit to accurately capture your story."*

#### 3. Collaboratively set the Agenda

**Why?** If you and the patient have built the agenda together, you are both working on a successful outcome. Time efficiency is improved when an agreement has been made on what will be covered.

**What?** Ask the patient what he or she wants to address, provide your agenda items, and then determine a mutually agreeable agenda for the visit?

**How?** *""What are you hoping we can address in today's visit?" (Wait for patient response) "What else?" (Gather the list) "In terms of what we'll cover together, I'd like to suggest that first I hear more about the difficulty you've been having, then I'll need to ask questions t o get a better idea of what is going on, and after that, we will work on next steps. How does that sound?"*

#### 4. Demonstrate Empathy verbally

**Why?** Empathic statements let the patient know we care. Empathic statements are therapeutic, improve outcomes and save time in a visit. Because we are not in the same physical location as the patient, these statements highlight our humanity.

**What?** An empathic statement is a statement that addresses the emotion a patient has expressed or may be feeling.

**How ?** *"I can only imagine how difficult this must be for you." "I'm here to help you through this." I wish I could be there with you in person." "I hear worry in your voice." "I can hear how hard this has been on you." "I'm excited about your progress too." "I would feel frustrated as well." "It sounds like you have had some very difficult days recently."*

#### 5. Elicit the patient narrative of the History of Present Illness

**Why?** Allow patients to feel heard while also providing valuable insight for improved diagnostic accuracy.

**What?** Allow patients to tell their story, in their own words.

**How?** *"Tell me more about your* (chief concern, worry, etc.)."

#### 6. Engage in Reflective Listening

**Why?** Patients don't know what we hear and understand unless we repeat back to them what they have just told us. Repeating back the story helps both the clinician and the patient move forward.

**What?** Reflective listening is a summary of the key points that a patient has just expressed.

**How?** *"It sounds like…" or "If I'm hearing you correctly…" or "Let me reflect back that key points you've shared…"*

#### 7. Share diagnosis and information in the context of the patient's perspective

**Why?** Patients learn best when new information impacts something that personally matters to them. Patients are also more motivated for behavior change when they are aware of how it will benefit something that is important to them.

**What?** Identify what is most important to the patient, such as the biggest concerns or goal. Then identify how any diagnosis or information and treatment planning might impact what matters most to the patient.

**How?** *"You have had several low blood sugar events that you can't explain that have occurred this past week." Or "It looks like your son has eye irritation and not pink eye. That means he is not contagious and can go to the birthday party."*

#### 8. Collaboratively develop the treatment plan

**Why?** Patients will be more motivate and confident in their capacity to manage their health, leading to improved clinical health outcomes.

**What?** Provide sufficient information to patients, invite them to share their ideas and preferences, and then incorporate them into the plan.

**How?** *"I am glad you made a virtual visit appointment, so we can discuss your low blood sugars. There are a number of things you can do to prevent a low blood sugar. First, I would recommend checking your blood sugar more frequently for the next week so that we can get a clearer picture of what is happening. Are you willing to try that?"* Or *"Strategies to treat eye irritation can include eye drops or just watching and waiting. It is important for your son to wash his hands more frequently and to avoid rubbing his eye, although that is easier said than done. Eye drops may help your son's eye look better, although they are not necessary. Would you like to talk more about eye drops?"*

#### 9. Have the patient repeat back what they understand

**Why?** Asking patients to repeat what they understand provides an opportunity to correct any misunderstanding or fill in any gaps before the visit ends. It also helps patients recall the information after the visit, and thus, facilitates their health management.

**What?** Also called teach back, it is the process of asking patients to restate what they understand and what they are going to do next.

**How?** *"To make sure my recommendations made sense, will you tell me what you heard are the next steps?"*

#### 10. Provide Closure

**Why?** Patients will look to you for a sign that the work of the visit is done. Since it is an expected practice in relationships, providing close also reinforces the personal connection you have with them.

**What?** Give a clear signal to the patient that the visit is coming to a close.

**How?** "*It's time to wrap-up our visit for today. I'm so thankful that you didn't wait to share this concern. I will put a note in your chart. I hope you had a good visit and will consider another virtual visit in the future."*



© The Cleveland Clinic Foundation. All Rights Reserved.
