## Supplementary material for "Implementing a Telemedicine Curriculum for Internal Medicine Residents during a Pandemic: The Cleveland Clinic Experience": Precepting Guide for Telemedicine at CCF

**Before your clinic half day**

**Review** resident schedule. This will help find any discrepancies & assist with the overall flow of the clinic (e.g. Recognizing high risk patients, complex medical decision making, patients that are not the PCP residents for communication and follow up purposes, etc.)

**On your clinic half day**

**Huddle** with resident team **at the start of clinic** to go through work flow, **assignment of where residents will be making their telemedicine/virtual visits,** answer questions, set expectations, and to determine potential patients in which you can use the exception rule***. **Include medical assistant(s)in the huddle so that** communication with team & identify modality preferred such as call/text/page

**Review Assignment or Decide** which room each resident will be using for their telemedicine phone calls or any video based virtual visits (Am Well App, FaceTime, Google Duo)

*****GE modifier**, PC exception: preceptor available during visit but not physically participating; (encounter not observed, no direct interaction with patient); NOTE: can be used for all PGY levels at the discretion of the preceptor; sign note using “.pcexception” Billing can be no more than **LEVEL 3**

**GC modifier**, direct precepting at time of visit through direct observation of care and bidirectional communication with patient, includes completion of preceptor participation prior to end of visit

**Set expectations** with resident team at the start of clinic about communication and use of PCP flex time:

**PCP Flex Time** can be used for the following:

1) In-basket: Review the resident’s in-basket and that of their attached in-basket team members
2) Schedule: Print and review their patient schedule for their next Y week in 5 weeks to provide rescheduling/conversion information. Schedule given to appropriate team member based on site. (G10: give to Monice McQueen to assign to her team/STJ: use the dot system on EPIC)
3) Panel Management: Review their LCC patient list with attention to the following

Update Care Team if needed. e.g. Staff PCP name should be only one of their care team preceptors

Care gap registry to review uncontrolled diabetic patients, etc

Review and update their patients’ problem list

4) End of residency requirements: **PGY3’s will be working on their patient panel sign out list for incoming interns in July**. At least one faculty member from each team should review the list with particular attention to review of high risk patients.

**At the end of your clinic half day**

Ask for feedback from all team members

Provide resident(s) feedback on their telemedicine/virtual visits

Have residents forward any telephone/virtual encounters to you for co-signature/addenda

**Telemedicine phone encounters**

1. The resident will start the phone encounter by the established time. **next page w/details/pics** Follow protocol per clinic site (e.g. G10: If they reach the patient the dot must be turned **blue**)
2. Resident will introduce themselves and identify the resident clinic supervising physician at initiation of call. They must verify patient identity and location.
3. Resident will start documentation (we suggest using .TAMVIRTUALVISIT for template) and evaluation of patient as they would in a face to face visit
4. Resident will need to specify amount of time spent on the phone with the patient at the end of the visit along with the follow up plan for the patient.
5. If the visit was deemed appropriate for PC exception modifier during the initial huddle 🡪 resident will complete telephone encounter and then present to the preceptor pertinent details of the encounter with attention to the care plan they provided the patient. **IF NEEDED** Residents can re-contact the patient by phone if the plan needs modification after precepting. They are also encouraged to put the patient on hold if an unexpected concern arises during the phone encounter so they can discuss with their preceptor.
6. Resident will route the encounter to staff for billing and this will be billed with modifier **GE-there has been minimal guidance on this due to uncharted waters. At this time bill the time you spent on the encounter with resident in review and document this time specifically.**

***NOTE: Although you may have deemed a visit to be PC exception, the resident should be informed that if they have any questions or encounter any obstacles during the encounter, they can put the patient on hold and ask the preceptor. If a therapeutic plan is discussed please follow billing instructions for GC modifier below:***

7. If using GC modifier—visit discussed and precepted during the visit time🡪 resident will start the encounter and once completed will ask the patient to “hold while plan is being discussed with Dr. X.”

Preceptor and resident then work on finalizing the therapeutic plan and staff will participate/be present for plan discussion or additional care. Resident will then return to the line and say “I have discussed the plan with Dr. X …”

8. AT CONCLUSION of the phone call and precepting, follow protocol per clinic site
9. **Copy of the note must be sent to the PCP if the resident is taking care of a patient for whom they are not the PCP resident; identify these during the review and huddle stages**)

Documentation: **refer to 4C communication and State of Ohio specific requirements regarding documentation of an e-visit/ telephone visit.**

**Please ask all residents to have this documented in their encounter**

*“This telehealth encounter is provided under a state of emergency due to COVID19 and is for care for condition where providing the care is supportive of minimizing potential exposure and/or transmission of COVID19”*

The scheduled phone calls now open a template that mimics an office visit template (as opposed to a telephone encounter). With the change:

- A reason for the visit is **no longer required**.
- Continue adding “.TAM” at the beginning of the note, and selecting phone encounter.
- Exiting pre-chart view is now required (see below)
- A **chief complaint is now required** in order to close the chart (see below)
- Charge capture is needed before signing the chart (see below)
- The **blue dot** system remains in place (for checking in and out when you reached the patient)
- Please add a line on “Time spent” at the end of the note (example: “Time spent was 13 minutes”)

**Step by step:**

When you first open the chart, you need to “start your visit” (get out of pre-chart mode). This can be done by clicking on Start visit as the image shows:

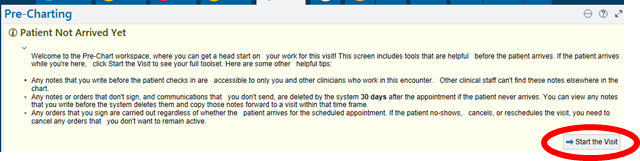

Then, go to Intake  🡪
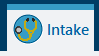

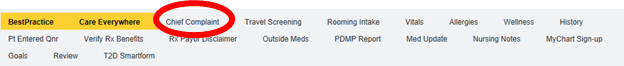
Then, add a chief complaint:

The rest of the visit works like it did before.

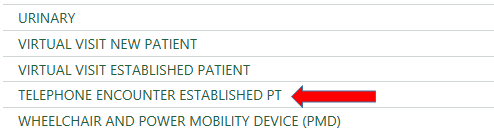
For charge capture, scroll down through the options and select Telephone encounter:

And select from the three options highlighted below in red (time-based)

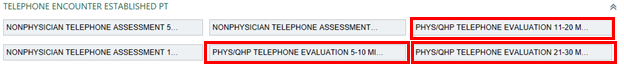

**Virtual Visit encounters**

Residents will be slowly on-boarded to virtual visits. Since this is a new platform for them, recommend discussing each visit with them as indicated below.

1. Resident will start the virtual visit via American Well app or FaceTime. They must use a patient examination room for the duration of the visit. Resident will introduce themselves and identify the resident clinic supervising physician for the virtual visit. It is recommended resident explains that he/she will step out to precept after the initial part of the visit.
2. They must document patient’s willingness to participate in a virtual visit and verify patient identity and **document** location (i.e. patient in Cleveland, OH).
3. Visit completed in entirety (residents have been given tool for physical exam evaluation for virtual patients); NOTE sensitive exams MUST be CHAPERONED just as if the patient were in the clinic.
4. Resident will inform patient that they will return after discussing with the preceptor.
5. Resident will step out and discuss plan with preceptor
6. Both resident and preceptor will enter the examination room to finalize the plan with the patient on the virtual platform regardless of PGY status.
7. Resident will route the encounter to staff to close the encounter and bill with inclusion modifier **GC**
8. AT CONCLUSION of the phone call and precepting, follow protocol per clinic site (e.g. for G10: If they reached the patient the dot was turned **blue** and then it is routed to their scheduling POD pool to “check in and check out” the patient.)
9. Note must be CC’d to the PCP if the resident is taking care of a patient for whom they are not the PCP resident; identify these during the review and huddle stages)

Documentation: **refer to 4C communication and State of Ohio specific requirements regarding documentation of virtual visits.**

**Please ask all residents to have this documented in their encounter**

*“This telehealth encounter is provided under a state of emergency due to COVID19 and is for care for condition where providing the care is supportive of minimizing potential exposure and/or transmission of COVID19”*

*This is a fluid process. Changes will happen as new processes unfold and critical to this is an iterative process so PLEASE send us your feedback and suggestions!*

**NOTE:** Due to the time sensitive nature of these changes, LCC templates have not yet been modified to optimize the flow. We are looking to your feedback to best handle this important aspect of the process with attention to access and educational opportunity as both remain top priorities.
