## Supplementary material for "Implementing a Telemedicine Curriculum for Internal Medicine Residents during a Pandemic: The Cleveland Clinic Experience": Telemedicine Survey - Pre-Participation

### Perspectives on Telemedicine

* Required

1. Current PGY year: *

*Mark only one oval.*

- ( ) PGY1
- ( ) PGY2
- ( ) PGY3

2. Prior experience with telemedicine: *

*Check all that apply.*

- [ ] Medical school course
- [ ] Telemedicine demonstration by an attending
- [ ] Note to date

Other: [ ] ______________________

3. I feel comfortable conducting a telemedicine visit. *

*Mark only one oval.*

- ( ) Very uncomfortable
- ( ) Uncomfortable
- ( ) Neutral
- ( ) Comfortable
- ( ) Very comfortable

4. What benefits do you see to using telemedicine? (check all that apply) *

*Check all that apply.*

- [ ] Improved patient outcomes
- [ ] Stronger physician - patient relationship
- [ ] Access to data (e.g. home measurements) or resources (e.g. consultants)
- [ ] Improved efficiency
- [ ] Better access for patients

Other: [ ] ____________________

*Skip to question 5*

5. What barriers do you see to using telemedicine? (check all that apply) *

*Check all that apply.*

- [ ] Providers lack of familiarity with the technology
- [ ] Weaker physician - patient relationship
- [ ] Too difficult for my patients to use
- [ ] Lack of time to implement care
- [ ] Privacy concerns
- [ ] Reimbursement concerns
- [ ] Documentation concerns

Other: [ ] ____________________

6. I expect to integrate telemedicine into my future practice. *

*Mark only one oval.*

- ( ) Strongly disagree
- ( ) Disagree
- ( ) Neutral
- ( ) Agree
- ( ) Strongly agree

*Skip to question 7*

7. Any other comments?

*Skip to question 8*

8. I have started conducting virtual visits (phone call or video) in my clinic and I am interested in participating in a 30min focus group. *

*Check all that apply.*

- [ ] I have not started virtual visits yet
- [ ] No
- [ ] Yes (type email below in "other")

Other: [ ] ____________________

This content is neither created nor endorsed by Google.

Google Forms
