## Supplementary material for "Implementing a Telemedicine Curriculum for Internal Medicine Residents during a Pandemic: The Cleveland Clinic Experience": Telemedicine Survey - Post-Participation

### Telemedicine Program Evaluation

* Required

1. Current PGY year:

*Mark only one oval.*

- PGY1
- PGY2
- PGY3

2. Overall, I was satisfied with this learning activity.

*Mark only one oval.*

- Strongly disagree
- Disagree
- Neutral
- Agree
- Strongly agree

3. I would recommend this learning activity to others.

*Mark only one oval.*

- Strongly disagree
- Disagree
- Neutral
- Agree
- Strongly Agree

4. The learning activities and/or materials were effective in helping me learn the content. *

*Mark only one oval.*

- Strongly disagree
- Disagree
- Neutral
- Agree
- Strongly agree
- Other: ______________________

5. I learned new knowledge and skills from the telemedicine curriculum. *

*Mark only one oval.*

- Strongly disagree
- Disagree
- Neutral
- Agree
- Strongly agree

6. The scope of the material was appropriate for my needs. *

*Mark only one oval.*

- Strongly disagree
- Disagree
- Neutral
- Agree
- Strongly agree

7. I found the material in this learning activity to be relevant and up-to-date. *

*Mark only one oval.*

- Strongly disagree
- Disagree
- Neutral
- Agree
- Strongly agree

8. The content was relevant to my job-related needs. *

*Mark only one oval.*

- Strongly disagree
- Disagree
- Neutral
- Agree
- Strongly agree

9. I will be able to apply the knowledge and skills learned for future encounters. *

*Mark only one oval.*

- Strongly disagree
- Disagree
- Neutral
- Agree
- Strongly agree

10. What about this learning activity was most useful to you? *

11. What about this learning activity was least useful to you? *

12. Please provide any additional comments or recommendations concerning this program. *

This content is neither created nor endorsed by Google.

Google Forms
